# Deep-Learning Model for Personalized Prediction of Positive MRSA Culture Results Using Patient’s Time-Series Electronic Health Records

**DOI:** 10.1101/2023.06.08.23291072

**Authors:** Masayuki Nigo, Laila Rasmy, Ziqian Xie, Bijun Sai Kannadath, Degui Zhi

**Author notes:** **Corresponding Author:** Masayuki Nigo, MD, Division of Infectious Diseases, Department of Medicine, Houston Methodist Hospital, Texas Medical Center, **Address:** Scurlock Tower, Suite 1512, 6560 Fannin Street, Houston Tx, 77030, **Tele:**, **Email:**.

## Abstract

Methicillin-resistant Staphylococcus aureus (MRSA) is a common bacterial cause of morbidity and mortality. Our deep-learning model (PyTorch_EHR) processes time-series structured electronic health record (EHR) data, including previous cultures and antimicrobial exposures, to predict the lab result of MRSA culture positivity over the next two weeks. After training and evaluation on data from 8,164 MRSA and 22,563 non-MRSA patient events from Memorial Hermann Hospital System, Houston, Texas, the PyTorch_EHR outperformed traditional machine learning methods logistic regression and light GBM (Area Under the Curve of Receiver Operating Curve [AUC]^PyTorch_EHR^=91.12%, AUC^LR^=85.91%, AUC^LGBM^=89.11%). External validation using the MIMIC-IV dataset of 393,713 patient events from a tertiary care center in Boston, Massachusetts, confirmed PyTorch_EHR’s accuracy (AUC^PyTorch_EHR^=85.50%, AUC^LR^=83.24%, AUC^LGBM^=82.48%). The model maintained its accuracy across most subgroup analyses based on infection type. The cumulative incidence curves based on our model successfully high-, medium-, and low-risk patients. This study demonstrates the potential of deep-learning models to predict the presence of MRSA-positive cultures to optimize MRSA antimicrobial therapy.

## Background

Methicillin-Resistant *Staphylococcus aureus* (MRSA) is one of the common pathogens causing both hospital-acquired and community-associated infections.^1^ Since this unique pathogen eliminates the majority of beta-lactam class antibiotics as a treatment option, physicians often need to add an antibiotic, such as vancomycin, to empirically treat this pathogen when suspected. Considering the side effect profile of vancomycin and antibiotic stewardship standpoint, it is highly desirable to avoid unnecessary antimicrobial therapy.^2^ Furthermore, a recent study showed the absolute benefit of empirical therapy against MRSA is 0.1% or less.^3^ Therefore, accurately identifying those high risk patients is critical to avoid unnecessary side effects from the empirical therapy with preserving the benefit of treatment. Though multiple clinical factors have been proposed as risk factors for MRSA infection, they have multiple limitations. ^4,5^ Commonly, the tested population is limited to specific populations, such as patients with ventilator-associated pneumonia.^6^ Due to the complex association of each risk factor, it is often difficult to discern actual risks when multiple risk factors simultaneously exist. For example, previous exposure to cephalosporine and fluoroquinolone were considered risk factors.^7,8^ The risk seems to accumulate when multiple antibiotics were previously prescribed.^9^ Further, the optimal timeline between the index infection and the presence of the risk factor is not well established, and often arbitrary duration is used.^10^ More flexible models which can integrate multiple risk factors and various timing of the risk factors are warranted for frontline physicians to safely decide the necessity of empirical antibiotic therapy.

Electronic medical records (EHR) became widely available in the U.S. since the Meaningful Use program was introduced as part of the 2009 Health Information Technology for Economic and Clinical Health Act. EHR became a rich data source for daily clinical practice and research purposes. The more data in EHR expands, the more information become available at physicians’ disposal to process and interpret to determine the patients’ management. Artificial intelligence (A.I.) has become an attractive technology to process real-time big EHR data in health care and achieve personalized medicine by processing a wide range of data. A.I. could reveal complex relationships among numerous factors in EHRs. Additionally, A.I. has been applied to various types of data, such as genetic, image, and EHR data.^11,12^ However, a few models currently predict drug-resistant bacteria.^13–15^ Even those available models only use very limited input data, such as basic demographics and previous susceptibility results or a limited number of patients.^13^ Furthermore, the model only predicts the index culture or screening, which may not be optimal in clinical use to guide antibiotic therapy.^16^ Deep-leaning-based models, such as recurrent neural network (RNN) models, have a significant advantage in time sequence events because the fundamental model structure allows sequential inputs into the model. Further, RNNs with medical code embedding can take inputs directly from a real-time EHR data stream, automatically adjust to reflect subtle changes, and provide real-time outputs. PyTorch_EHR has been successfully applied to predict various clinical outcomes. ^14–16^ Despite the potentially high expressive power of deep learning models, no deep learning models using EHR data for predicting MRSA cultures are available.

This study aims to create a deep-learning-based prediction model for positive MRSA culture using big time-series electronic health record data and compare the traditional machine learning models. Further, we evaluated the model’s generalizability in external EHR data in a different region in the U.S.

## Method

Two datasets were used in this study; Memorial Hermann Hospital System (MHHS) and Medical Information Mart for Intensive Care (MIMIC) - IV for model training and external validation. Patient data were retrospectively retrieved from the microbiology database at Memorial Hermann Hospital System, Houston, Texas. MIMIC-IV ver. 2.1 is a relational de-identified EHR database containing real hospital encounters from a tertiary academic medical center in Boston, MA, USA.^20^

In MHH datasets, patients aged more than 18 years old were obtained from 1/2018 and 4/2021. Those patients had at least bacterial cultures during the study period. To avoid an imbalanced dataset, we randomly selected approximately 5000 patients with MRSA, non-MRSA positive cultures, including MSSA and other types of bacteria, and negative cultures. Demographic data, admission data, diagnostic & procedure codes, antibiotic administration, other ID related test results, and previous microbiological data, including the type of cultures, name of bacteria, and all antibiotic sensitivities, are obtained from the database. The admission ward information was converted only to emergency department (ED), intensive care unit (ICU), and intermediate unit (IMU) so that that information can be generalizable to MIMIC-IV data. Microbiology table includes cultures and other infectious diseases tests, such as serologies. To avoid any label leakage, we used only results reported by the index time. The laboratory orders were included without results if they were ordered by the index time. For diagnostic and procedure codes, ICD-9 or 10 codes were used. Since other data tables, such as antibiotics, did not contain standardized codes for medications, free text, such as “vancomycin”, was used. Extracted data were cleaned and converted to categorical data to fit Pytorch_EHR scheme.

Regarding MIMIC-IV data, all patients aged more than 18 years old who had bacterial cultures were similarly retrieved. To validate the generalizability of the model, each data table was mapped with the MHHS data table. Only data mapped with MMHS data were used in MIMIC-IV dataset. Since MIMIC-IV datasets aggregated the ICD and procedure codes at each encounter level, we only used the ICD or procedure codes only reported in the previous encounters to avoid label leakage. Microbiology event table was used to identify the patient. A total of 25,599 *S. aureus* from various cultures were found in the table. 19,605 isolates (76.6 %) were tested for antimicrobial sensitivity for various reasons; multiple positive cultures with *S. aureus* in a short period and positive wound cultures due to multiple organisms. After removing positive *S. aureus* within seven days after positive MSSA or MRSA, only *519 S. aureus* isolates did not have any recent sensitivity to classify MRSA or MSSA. Those isolates were classified into the negative MRSA group. We further divided the datasets as 70:10:20 to fine-tune the pre-trained model with MHHS datasets.

In terms of subgroup analysis, we used ICD codes to identify the patients with the ICD code within the two weeks period in the MMHS dataset. Since MIMIC-IV only provided ICD codes at the encounter levels, we used the encounter to find the patients with the ICD codes within the encounter.

### PyTorch_EHR Prediction Model Scheme

We set a two-week window for the prediction, and any first culture within the window was used as an index culture (Figure 1). Considering the majority of MRSA infections or new infections are diagnosed within two weeks, we decided to use a two-week window. This prediction window allows not only predicting at the time of culture but also cultures obtained after initiation of empirical antibiotics, which is essential for physicians to decide whether empirically start or continue MRSA antibiotic coverage. Some patients had multiple cultures over time, including both positive MRSA and non-MRSA cultures. Those patients can be included in both MRSA and non-MRSA groups, depending on the type of positive or negative culture the patient had during the window period.

**Figure 1.**
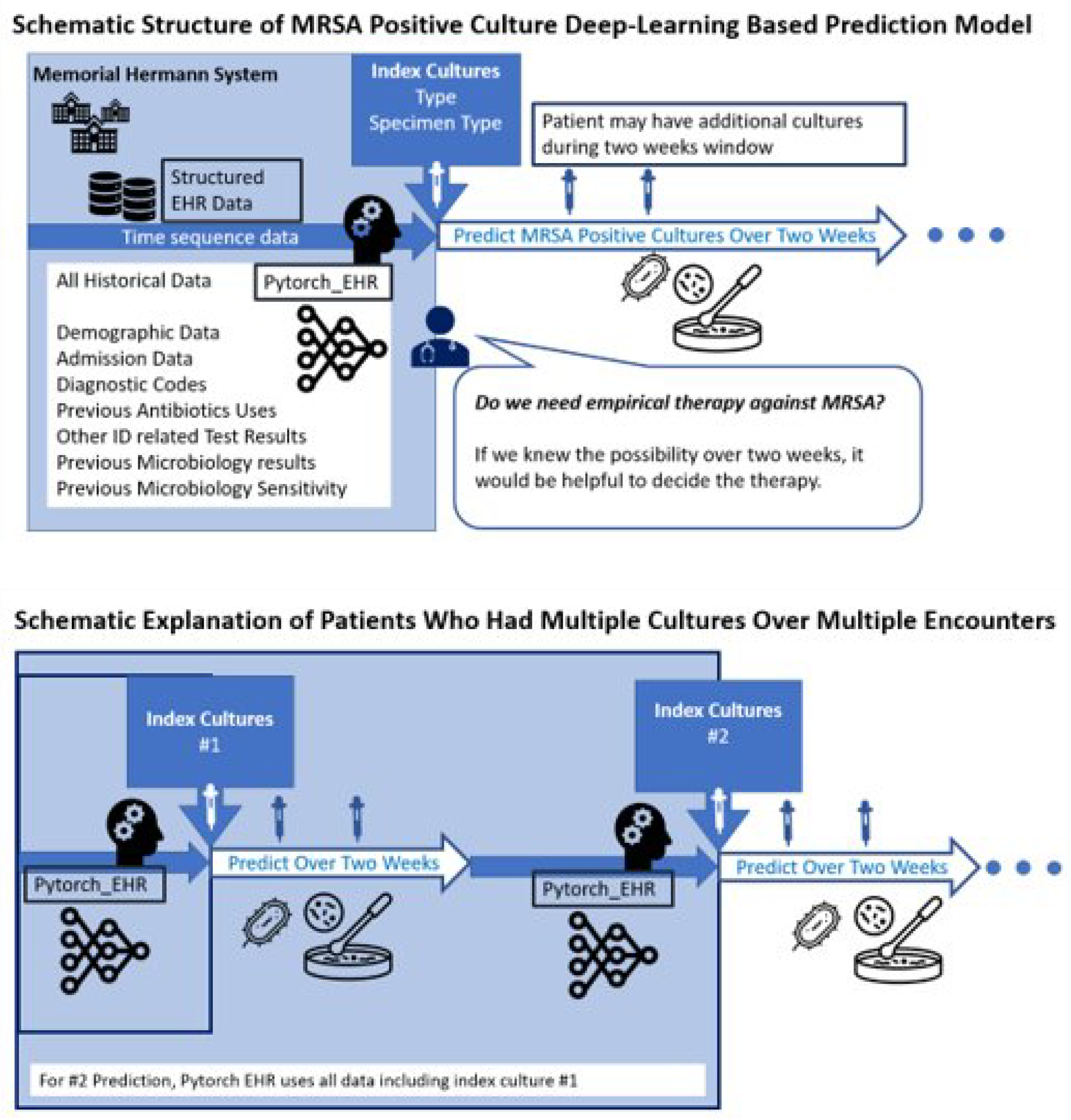
Schematic Structure of Deep-Learning Based Prediction Model for Positive MRSA Cultures.

In terms of the deep-learning platform, we use Pytorch_EHR for clinical outcomes predictions using categorical data from electronic medical records. Pytorch_EHR implemented a model of recurrent neural networks (RNN). We choose the gated recurrent unit (GRU) RNN architecture, which is known for being an efficient sequential deep learning architecture for clinical event predictions. (See S Figure 1) The source code of this model is publicly available to enable its application and further evaluation by other researchers.^21^ In addition to categorical data, Pytorch_EHR handles the time difference between visits for a better temporal representation of patient history to improve accuracy. (See S Figure 2)^22,23^ In the project, we converted the interval to days from visits to accommodate the predictions for more acute issues.

**Figure 2:**
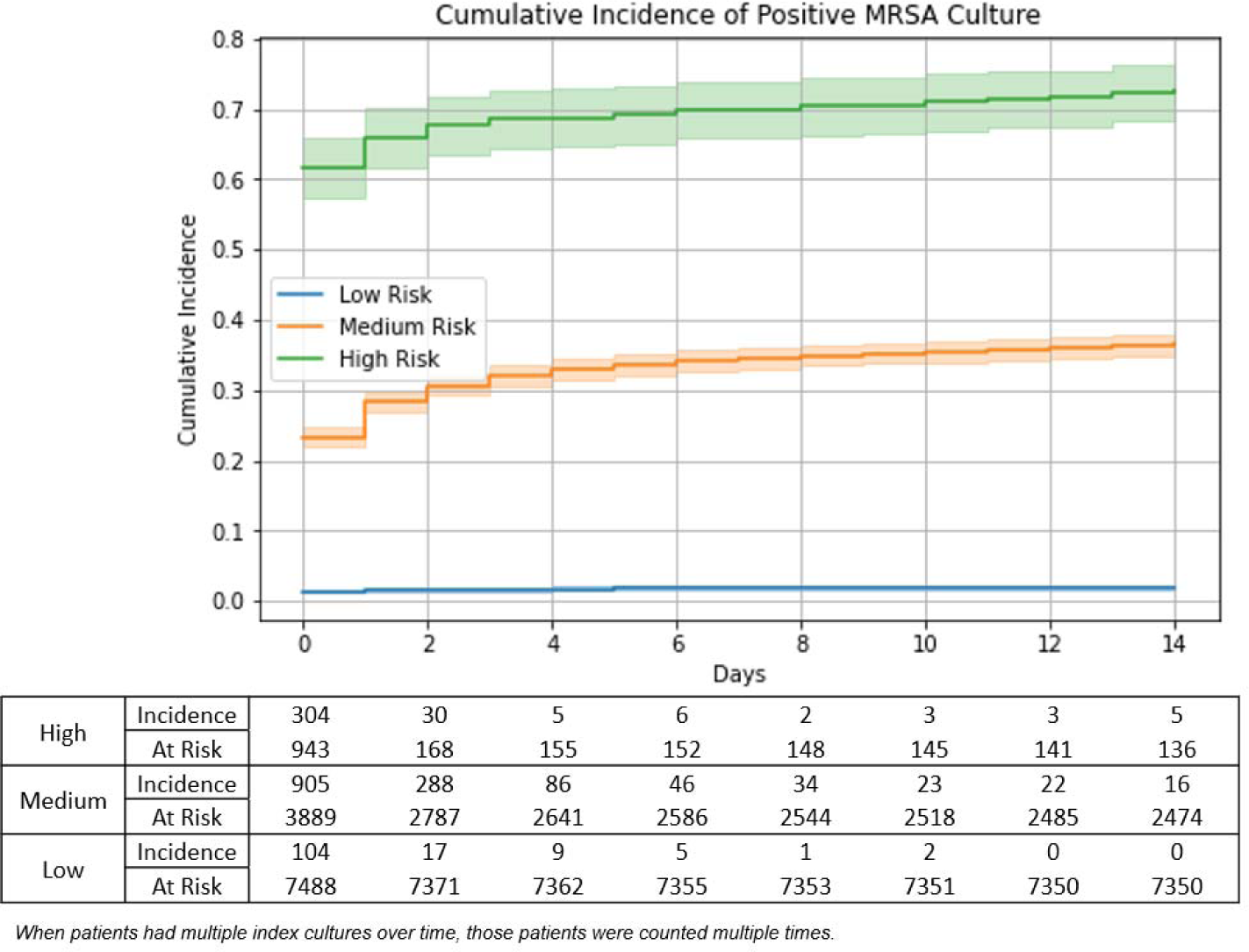
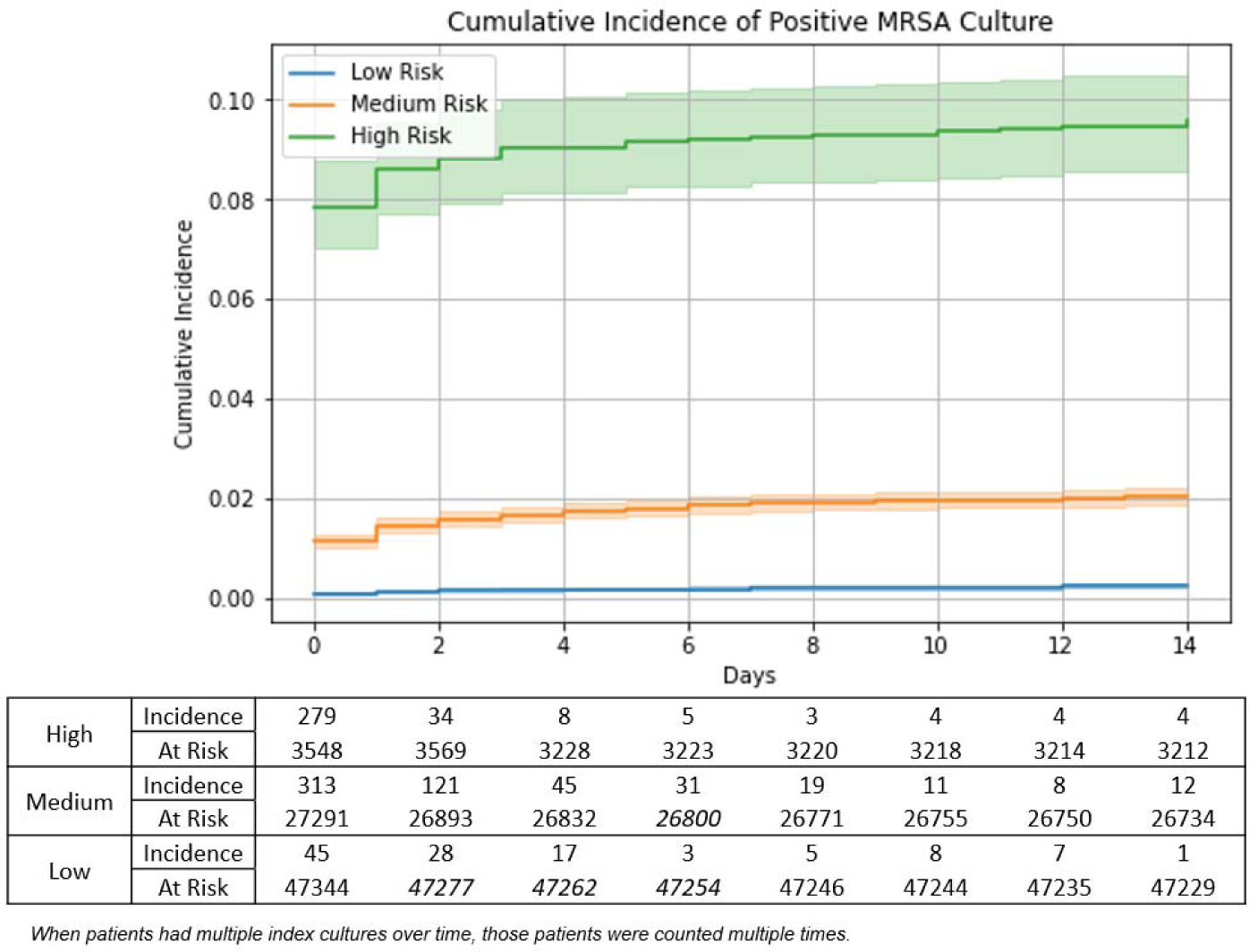
Cumulative Incidence Curve Over Two Weeks in MHHS and MIMIC-IV Datasets

For binary classification tasks, we compared our model against traditional machine learning algorithms: logistic regression^24^ and light gradient boost machine.^25^ For survival prediction, we used the DeepSurv^26^ architecture while replacing the multiple-layer perceptron layers with GRU layers for better sequential information modeling, similar to the way we modeled COVID-19 outcome prediction.^18^ Pytorch ver. 1.7.1 and Sklearn ver. 0.24.2 are used. After hyperparameter tuning for each model, we run ten times for each model training and test datasets to obtain the average of our model performance and confidence intervals.

This study is approved by Institutional Review Boards at the University of Texas Health Science Center at Houston and Memorial Hermann Hospital System.

### Model interpretation

For the interpretation of MRSA predictions, we used the integrated gradient technique^27^ to expose the factors that contribute to the personalized model predictions. For recurrent neural network-based models, we can achieve a more personalized explanation that shows the contribution scores for each clinical event at each patient day in the patient trajectory. This is unlike the standard population-level feature contributions that we can infer from logistic regression coefficients or light gradient boost machine feature importance scores. To evaluate our RNN-based model explainability, we reviewed the calculated contribution scores for each clinical event in the input of 10 patients. For facilitation, we visualized the contribution score per patient through a Tableau interactive dashboard, where clinicians can navigate different clinical events within different categories and along different visits within the patient history. This study was approved by internal review boards (IRBs) of the University of Texas Health Science Center at Houston and the Memorial Hermann Hospital System.

## Results

A total of 30,727 and 156,113 patients were identified in the MHHS and MIMIC-IV database, respectively. As described under Methods, a total of 30,727 patients in MHHS database were randomly selected among them. The aggregated patient characteristics are described in table 1. Since some patients were classified into MRSA and MSSA groups, the total number of patients in both groups was 30,727. The patient features were used once if the patient had two or more events in the same group. The last demographic features were used to describe the characteristics when patients are classified more than twice into one group. Overall, MRSA group had a higher number of ICU admission (4.3% vs 2.0% in MHHS and 31.7% vs. 16.7% in MIMIC-IV) and ED patients (66.4% vs. 36.5% in MHHS and 51.3% vs 35.0% in MIMIC-IV). As MIMIC-IV was originally developed based on ICU database, MIMIC-IV database included a higher number of ICU patients. Intermediate care unit (IMU) status was not included in the MIMIC-IV data. The most common age group was 55 - 65 years old in all groups. Gender was equally distributed in all groups. As expected, given the origin of data (MHHS for Houston and MIMIC for Boston), MHHS databases had more Hispanic compared to MIMIC IV (10.5 % vs. 3 – 4 %, respectively). Among races, Caucasian was most common in all groups. In terms of antibiotics administration, vancomycin is the most common antibiotic used followed by cefepime in MHHS. Ceftriaxone is the second most common antibiotic in MIMIC-IV database. Regarding the cultures, blood and urine cultures are common cultures taken during the study periods.

**Table 1.** Characteristics of Patients with Positive MRSA cultures and without Positive MRSA cultures^a^.

|  | Memorial Hermann Hospital System |  | MIMIC-IV |  |
| --- | --- | --- | --- | --- |
|  | MRSA group<br>N = 8164 | Non-MRSA group<br>N = 22,563 | MRSA group<br>N = 4107 | Non-MRSA group<br>N = 152,006 |
| **Hospital location<sup>b</sup>:**<br>**Top 5** | N (%) | N (%) | N (%) | N (%) |
| ICU | 354 (4.3%) | 166 (2.0%) | 1302 (31.7%) | 25319 (16.7%) |
| IMU | 283 (3.5%) | 124 (1.5%) | - | - |
| ED | 5421 (66.4%) | 2981 (36.5%) | 2106 (51.3%) | 53185 (35.0%) |
| **Age** |  |  |  |  |
| 55 – 65 | 1965 (24.1%) | 3857 (17.2%) | 943 (23.0%) | 30196 (19.9%) |
| 65 – 75 | 1943 (23.8%) | 3637 (16.2%) | 934 (22.7%) | 28551 (18.8%) |
| 45 – 55 | 1483 (18.2%) | 2752 (12.3%) | 729 (17.8%) | 23956 (15.8%) |
| **Gender, Male** | 4200 (51.4%) | 9897 (44.2%) | 2338 (56.9%) | 86718 (57.0%) |
| **Ethnicity<sup>b</sup>** |  |  |  |  |
| Hispanic | 866 (10.6%) | 2358 (10.5%) | 159 (3.9%) | 5400 (3.6%) |
| Non-Hispanic | 5671 (69.5%) | 11210 (50.1%) | 2810 (68.4%) | 78559 (51.7%) |
| Unknown | 651 (8.0%) | 1064 (4.8%) |  |  |
| **Race<sup>b</sup>: Top 5** |  |  |  |  |
| White | 3123 (38.3%) | 6576 (29.4%) | 2153 (52.4%) | 55657 (36.6%) |
| African American | 1555 (19.0%) | 2878 (12.9%) | 406 (9.9%) | 12701 (8.4%) |
| Asian | 103 (1.3%) | 413 (1.8%) | 53 (1.3%) | 3233 (2.1%) |
| Other | 2174 (26.6%) | 4470 (20.0%) | 281 (6.8%) | 9872 (6.5%) |
| Unknown | 470 (5.8%) | 905 (4.0%) | 185 (4.5%) | 4991 (3.3%) |
| **Primary Language<sup>c</sup>** |  |  |  |  |
| English | 6314 (77.3%) | 12927 (57.7%) | 2699 (65.7%) | 74779 (49.2%) |
| Spanish | 248 (3.0%) | 743 (3.3%) | - | - |
| Unknown | 732 (9.0%) | 1260 (5.6%) |  |  |
| **Selected**<br>**Comorbidities<sup>d</sup>** |  |  |  |  |
| Cerebrovascular<br>accident | 918 (11.2%) | 4700 (21.0%) | 328 (8.0%) | 14360 (9.4%) |
| Congestive Heart<br>Failure | 1408 (17.2%) | 7021 (31.4%) | 1034 (25.2%) | 37401 (24.6%) |
| Chronic pulmonary<br>diseases | 1041 (12.8%) | 5779 (25.8%) | 1156 (28.1%) | 51492 (33.9%) |
| Cirrhosis | 149 (1.8%) | 1175 (5.2%) | 184 (4.5%) | 13126 (8.6%) |
| Chronic Kidney Disease | 1438 (17.6%) | 8049 (35.9%) | 1081 (26.3%) | 41702 (27.4%) |
| Hypertension | 3008 (36.8%) | 17429 (77.8%) | 2228 (54.2%) | 106436 (70.0%) |
| Diabetes Mellitus | 1790 (21.9%) | 9225 (41.2%) | 1411 (34.4%) | 52323 (34.4%) |
| Malignancy | 477 (5.8%) | 3380 (15.1%) | 767 (18.7%) | 45128 (29.7%) |
| HIV/AIDS | 136 (1.7%) | 541 (2.4%) | 56 (1.4%) | 2829 (1.9%) |
| **Total Antibiotics Given in The Group (days)<sup>b</sup>: Top 5** |  |  |  |  |
| Vancomycin | 28153 | 21185 | 7647 | 75879 |
| Cefepime | 19486 | 14687 | 3590 | 42530 |
| Meropenem | 11960 | 9542 | 1744 | 19448 |
| Ceftriaxone | 10674 | 8259 | 2879 | 49130 |
| Piperacillin-tazobactam | 10473 | 8227 | 2224 | 25558 |
| **Total Antibiotics Route Given in The Group (days)<sup>a</sup>: Top 5** |  |  |  |  |
| IV | 71749 | 54986 | 16160 | 191633 |
| PO | 13446 | 10317 | 10498 | 167895 |
| Enteric tube | 8275 | 6703 | - | - |
| Inhalation | 561 | 504 | 0 | 0 |
| Ophthalmic | 138 | 121 | 2 | 100 |
| **Total Culture Type Obtained in The Group<sup>b</sup>: Top 5** |  |  |  |  |
| Blood | 14741 | 33907 | 17557 | 128537 |
| Urine | 6263 | 29840 | 12402 | 215712 |
| Wound | 4769 | 7523 | 4424 | 11798 |
| Anaerobic | 2150 | 5966 | 5603 | 13214 |
| Body Fluid/Tissue | 1963 | 6443 | 1946 | 13974 |

a. When a patient had positive cultures for both MRSA and non-MRSA at different prediction periods, those patients were included in both groups.
b. Some patients had multiple encounters, orders, or documentation during study periods. Those are separately counted unless documented on the same date.
c. Some patients may speak more than one language. If multiple languages were documented, those are separately counted.
d. Since some patients had multiple encounters during the study period, diagnostic codes are summarized based on patient levels rather than encounter levels.

Table 2 summarizes the bacteria and diagnostic codes identified within the event periods. Of note, *S. aureus* and enterococci were the common bacteria in MRSA groups, whereas E. coli is the most common in Non-MRSA group. Bacteremia and skin soft tissue infection are more commonly seen in MRSA groups (8.0% in MHHS and 5.0% in MIMIC-IV). UTI was the most common diagnosis in non-MRSA groups (56.0% in MHHS and 13.8% in MIMIC-IV).

**Table 2:** Name of Bacteria Identified from Cultures and Type of Infections Based on ICD Codes

|  | Memorial Hermann Hospital System |  | MIMIC-IV |  |
| --- | --- | --- | --- | --- |
|  | MRSA group<br>N = 8164 | Non-MRSA group<br>N = 22,563 | MRSA group<br>N = 5789 | Non-MRSA group<br>N = 387,924 |
|  | N (%) | N (%) | N (%) | N (%) |
| **Bacteria Name <sup>b</sup>:** |  |  |  |  |
| 5 Common Bacteria |  |  |  |  |
| *S. aureus* | 8164 (100%) | 2117 (9.5%) | 5789 (100%) | 10724 (2.8%) |
| *Enterococcus spp.* | 848 (10.4%) | 3745 (16.7%) | 360 (6.2%) | 10140 (2.6%) |
| *E. coli* | 569 (7.0%) | 8028 (35.9%) | 287 (5.0%) | 27403 (7.1%) |
| *K. pneumoniae* | 373 (4.6%) | 2258 (10.1%) | 213 (3.7%) | 7619 (2.0%) |
| *P. aeruginosa* | 783 (9.6%) | 2224 (9.9%) | 463 (8.0%) | 5753 (1.5%) |
| **Types of Infections** |  |  |  |  |
| Sepsis | 1965 (24.1%) | 5621 (25.1%) | 411 (10.0%) | 8007 (5.3%) |
| Pneumonia | 1130 (13.8%) | 3882 (17.3%) | 675 (16.4%) | 14669 (9.7%) |
| Bacteremia | 657 (8.0%) | 1045 (4.7%) | 497 (12.1%) | 7531 (5.0%) |
| Skin Soft Tissue Infection | 2420 (29.6%) | 2721 (12.2%) | 762 (18.6%) | 10106 (6.6%) |
| UTI | 1151 (14.1%) | 12545 (56.0%) | 652 (15.9%) | 20929 (13.8%) |

Table 3 shows the prediction accuracy of the models. Deep-learning model, Pytorch_EHR model exhibited the highest Area Under the Curve of Receiver Operating Curve (AUROC) of 91.12% (91.023-91.214) (ROC curve: S Figure 3) compared to other machine learning models (LR 85.91% [85.911 – 85.912], and LGBM 88.51% [89.085 – 89.145]) in MMHS database. The findings were also consistently seen in MIMIC-IV database (Pytorch_EHR: 85.50%, LR: 83.24%, and LGBM 82.48%). We also evaluated the AUROC in each patient group with a specific diagnosis during the event. Although the AUC decreased by 5 - 10%, we had acceptable accuracy in each infection in MHHS database.

**Table 3.**
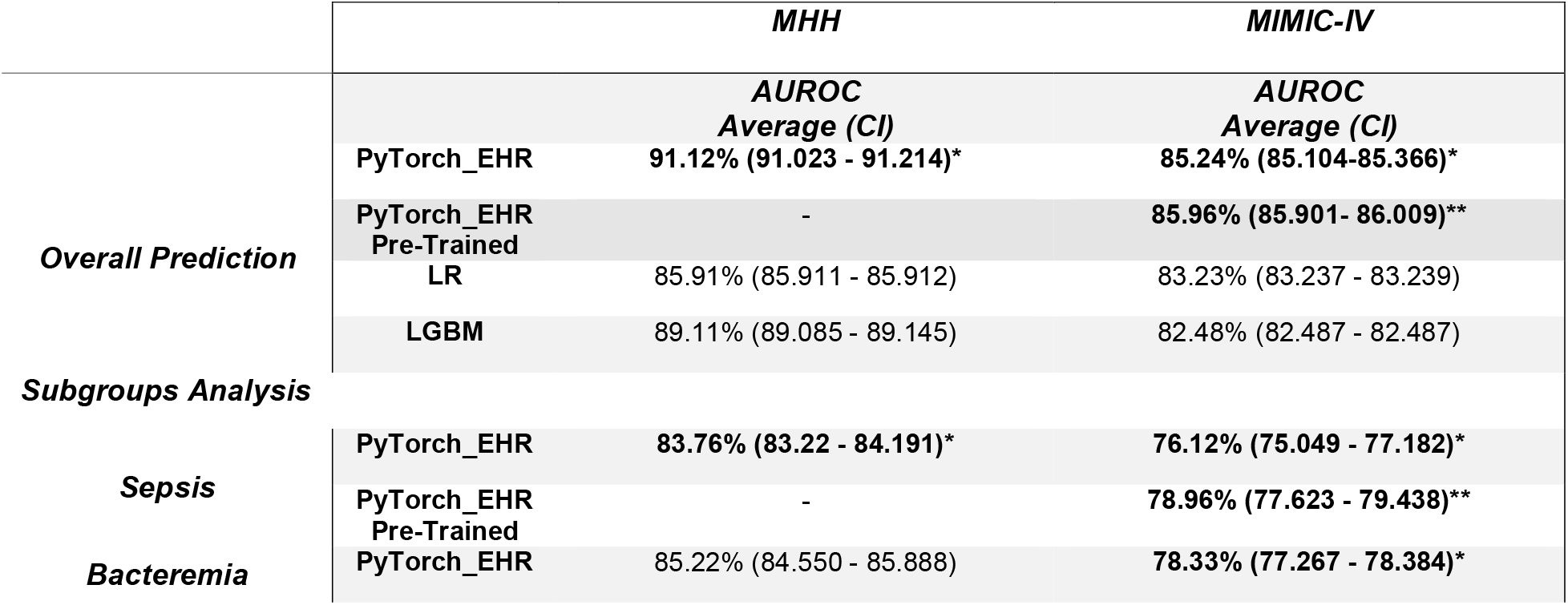
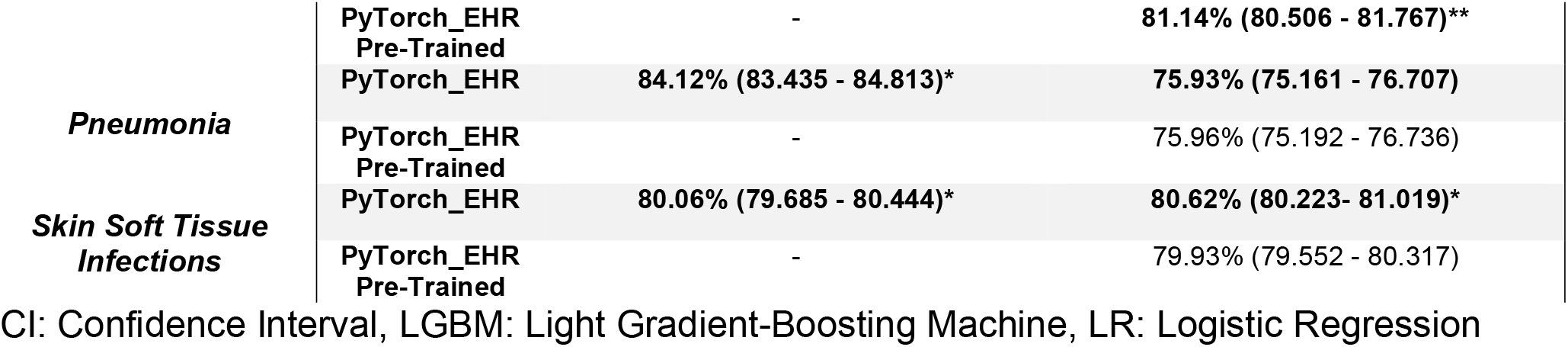
Outcome of Models in Overall and Subgroup Analyses

Figure 2 shows the cumulative incidence curve of MRSA culture over two weeks from the index culture. In both databases, our model nicely differentiated the patients with high and low risks of MRSA positive cultures. The cumulative incidence of MRSA positive culture in the MRSA group in MHHS database was 70.4%, whereas the incidence of in MIMIC-IV was approximately 11.9 %. The low incidence despite in high risk in MIMIC-IV was likely due to the overall incidence of positive MRSA culture in MIMIC-IV database.

Finally, we provide the visualization of the feature importance of example patients (S Figure 5). The degree of contribution of each feature is visualized as a bar graph. For example, the example patient #1 is a female patient aged between 45 – 54 years with multiple underlying multiple comorbidities listed on admission two days (−2 days) before the index culture (blood culture on index date). Our model identified a risk score of 0.541 (predicted as a positive patient). After the patient was admitted to the hospital, vancomycin and meropenem were initiated, and a blood culture was ordered. Subsequently, cultures identified MRSA over two weeks. We also visualized other example patients in S Figure 5. Of note, the same features may have different degrees of importance depending on the timing when the features are inputted to the model. In addition, the highly contributing features in this patient are not necessarily important features for other patients.

## Discussion

In this study, our deep-learning based MRSA predictive model outperformed other machine-learning models in both real-world MMHS and MIMIC-IV datasets. The model successfully “learned” patient-specific features to provide personalized risks of positive MRSA cultures over two weeks from index time. The model maintained better predictions even after transferring from MHHS dataset to MIMIC-IV dataset and tolerated the significantly imbalanced outcome in MIMIC-IV dataset. Compared to other existing models, our model successfully predicted the positive MRSA cultures not only on the index day but also over two weeks from the index day. This prediction window is more aligned with the daily clinical practice of physicians since physicians decide empirical antibiotic therapy to treat MRSA, such as intravenous vancomycin, not only for the culture of index day but also any subsequent cultures that may be related to the episode of infection after initiation of therapy. Our deep learning model takes the time sequence of the events in the patient history, which we believe is more consistent with the physician’s assessment in clinical practice. We also tested the model in various types of infection posing various MRSA risks, such as sepsis, bacteremia, and pneumonia. Although there were some decreases in the AUC, superiority and high performance are maintained, which supports this single model to be used in multiple types of infections.

Personalized medicine is of great interest in medical fields. Many studies in this matter focus more on genetic-based predictions rather than based on clinical data from electronic health records (EHR).^28^ EHR data have become a rich source of real-world data and provide invaluable information. Even without genetic data, we believe EHR data can be a great source for deep-learning models to achieve personalized medicine in multiple clinical settings. Furthermore, compared to traditional machine learning models, deep-learning can easily integrate time sequence data as inputs into the model, which provides significant advantages for those outcome predictions requiring sequential event inputs. Although Pytorch_EHR only uses categorical data from EHR, this model has provided high performance with advantages of relatively simpler preprocessing steps and flexible variable selections for input. This allows us to preserve model transferability and generalizability among different data sources.

Since MRSA emerged, multiple predictive models or risk factors for MRSA infections were proposed. Although the models provided various accuracy, the models often focus on a certain type of infection, such as pneumonia, to achieve and simplify the risk factors and models. Rhodes et al. used a machine learning model to predict community-acquired MRSA pneumonia.^29^ Although the time frame and patient population differed from our study, their model achieved AUC of 77.5%, which was a lower AUC than ours. Additionally, some risk factor-based models heavily rely on certain tests, such as the nasal MRSA PCR test from nare^30^, which hampers the model’s generalizability due to the tests’ availability and applicability to other types of infections. Further, some of the results may not be available at the time of starting antibiotics, which limits the usability of models in hospitals. On the contrary, our model carries a significant advantage since the model could take widely available data from EHR and can predict the outcomes even with some missing certain tests for the model. Our model can be used not only for the treatment decision but also, although the utility of contact precaution is still controversial, for Infection prevention to isolate the patients with high-risk groups in advance, even before culture results. Our model used a two-week time window to provide more meaningful predictions in clinical settings. Some predictive models only predict the index culture rather than overall risks.^16^ In order to be applied in the clinical setting, predicting a two-week window can be more impactful to clinicians when they choose antimicrobial therapy at the time of the initiation. Our cumulative incident curves based on our model prediction nicely differentiated the high and low-risk patients. The majority of patients had positive MRSA cultures on the index day, but approximately 15 % of high-risk patients had positive cultures after the index day, which could be missed if we only predicted the positivity of the index culture.

One of the challenges in deep-learning models is explainability of the models. As provided in S Figure 5, we visualized individual factors contributing to the model predictions. Since the model uses the sequence of time without dichotomizing the time frame with arbitrary cutoff, i.e., positive MRSA culture within 90 days, the contribution weight can be different depending on the patient and timing of events. Furthermore, those highly contributed events were not necessarily directly associated with the predictions of MRSA. The inputs could surrogate other underlying events. Caution is required to interpret the feature importance as those outputs can not be traditional risk factors we use in clinical settings.

There are multiple limitations in this study. First, due to the nature of retrospective studies, potential biases are not evitable. The findings in this study should be confirmed in prospective studies. In addition, although MHHS data are from Houston, Texas, and MIMIC IV is from Boston, Massachusetts, U.S., the model should be validated in different patient populations and high-risk populations, such as immunocompromised patients. Second, this model predicts positive MRSA cultures rather than infections. Since some patients may still have MRSA infections without positive cultures, the model should be used cautiously when there are significant concerns about MRSA when initiating antibiotics. Third, although we included multiple variables in the model, several important variables as known MRSA risk factors, such as residence in a long-term care facility, were not included. Furthermore, vital signs or other basic laboratory results were not included in this model. Those can be considered for future studies. Finally, although we showed the generalizability of the model in this study, the transferability of the model needs to be solved to use the deep-learning model widely.

## Conclusion

In this study, our deep-learning based predictive model successfully predicted positive MRSA culture over two weeks from index culture. Our study revealed the superiority against other traditional machine learning models in both MMHS and MIMIC-IV datasets with high performance, even in significantly imbalanced datasets and subgroup analyses. The model can be widely applied to various types of infections. Further studies in high-risk populations and prospective studies are warranted to confirm the findings.

## Data Availability

All data produced in the present study are available upon reasonable request to the authors

