## Supplemental File for "Deep-Learning Model for Personalized Prediction of Positive MRSA Culture Results Using Patient’s Time-Series Electronic Health Records"

**Supplementary Materials**

1. S Table 1: Features Included in this project
2. S Figure 1: Pytorch_EHR Pipeline
3. S Figure 2: Modified Data Structure of Pytorch_EHR
4. S Figure 3: Area Under Curve of Receiver Operating Characteristics
5. S Figure 4: Calibration Curve of Pytorch_EHR Model on Memorial Hermann System Data
6. S Table 2: Subgroup Analyses Results including Logistic Regressoin and Light GBM in MMHS and MIMIC-IV data
7. Features contributing to the individual prediction in an example patient
   1. **S Table 1: Features Included in This Project**

In this project, various tables/categories of structured electronic health records (EHR) data were used. This table summarized the features included in this project. Some features may be used with results when it was available before index time.

| **Category of Features** | **Features** |
| --- | --- |
| Demographics | Age^a^, Gender, Ethnicity, Race, Language |
| Encounter Information | Encounter location (ICU, ED, IMU), Admission diagnosis (String),  Duration of admission/encounter (if discharged before index time), |
| Diagnostic code | ICD10 or ICD9 codes |
| Procedure code | Procedure code (CPT codes) |
| Antimicrobials | Antibiotic name (string), route of the antibiotic, IV access |
| ID related test | HIV test, MRSA PCR, Legionella urine antigen, Treponemal antibody, etc. (150 different tests) |
| Cultures | Culture order, such as Blood culture, Culture source |
| Culture results | When the results were known before index time:  Positive, Negative, Isolate name |
| Sensitivity results | When the results were known before index time:  Name of bacteria, Name of tested antibiotic, Sensitivity interpretation of tested antibiotic against the isolate |

ICD: International Classification of Disease, ICU: Intensive Cardiac Unit, ID: Infectious Diseases, CPT: Current Procedural Terminology, ED: Emergency Department, IMU: Intermediate Unit, HIV: Human Immunodeficiency Virus, MRSA: Methicillin Resistant *Staphylococcus aureus*,

**S Figure 1: Pytorch_EHR Pipeline**

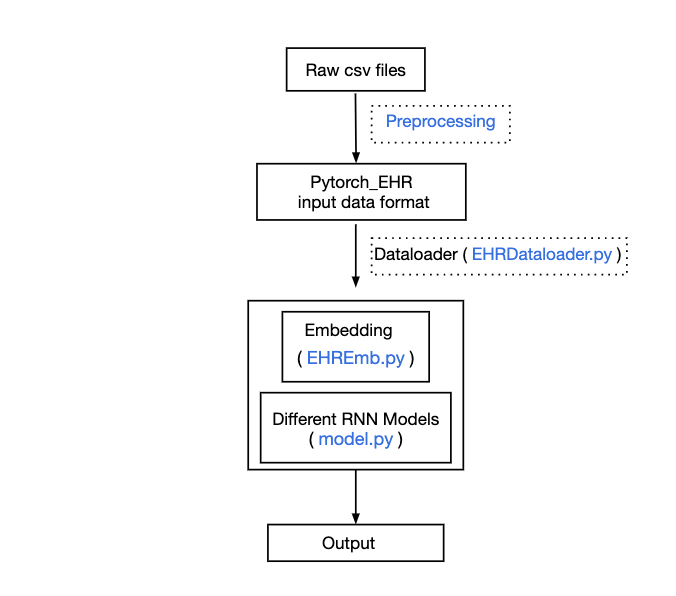

In this project, we used Pytorch_EHR scheme to predict positive MRSA cultures over two weeks from index culture. Original files were prepared as csv files for both Memorial Hermann Hospital System (MHHS) and MIMIC-IV. Data were preprocessed to be compatible to data format for Pytorch_EHR. In this project, all data were used as categorical data. The features were embedded and tested on different Recurrent neural network models and machine learning models.

**S Figure 2: Modified Data Structure of Pytorch_EHR**

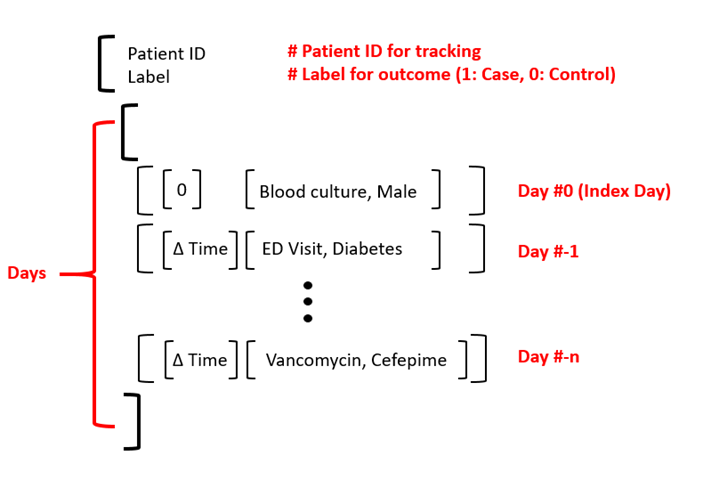

This is a data structure of Pytorch_EHR. Each patient data has patient ID and outcome label, followed by data inputs, including delta time between the days of inputs and inputs features. Input features can be multiple as any event happened on the day. Each feature was embedded before processing. During the training process, the model learns the relationship and timing of the features to predict the risk of positive MRSA cultures over two weeks.

**Figure S3.** Area Under Curve of Receiver Operating Characteristics

S3-1 Memorial Hermann Hospital System Data

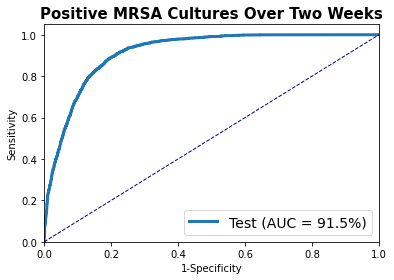

S3-2 MIMIC-IV Dataset

S Figure 4. Calibration Curve of Pytorch_EHR Model on Memorial Hermann System Data

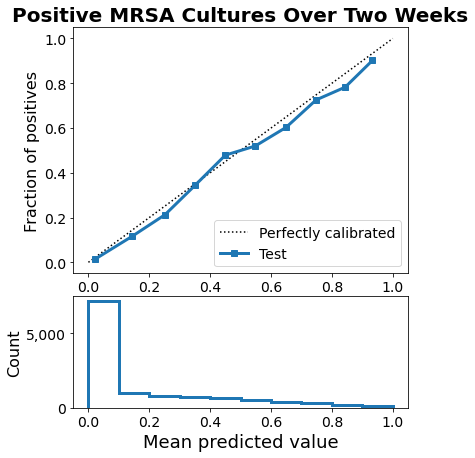

**S Table 2: Subgroup analysis results including Logistic Regression and Light GBM in MMHS and MIMIC-IV data**

| **Subgroups Analysis** |  | **MHHS**  ***Mean (CI)*** | **MIMIC-IV**  ***Mean (CI)*** |
| --- | --- | --- | --- |
| Sepsis | **LR** | 80.65% (80.644 - 80.649) | 68.83% (69.826 - 68.828) |
|  | **LGBM** | 82.90% (82.789 - 83.005) | 73.01% (72.603 - 73.407) |
|  | **PyTorch_EHR** | **83.76% (83.322 - 84.191)*** | **76.12% (75.049 - 77.182)*** |
|  | **PyTorch_EHR**  **Pre-Trained** | - | **78.96% (77.623 - 79.438)**** |
| Bacteremia | **LR** | 80.11 (80.111 - 80.118) | 71.11% (71.112 - 71.114) |
|  | **LGBM** | 85.08 (84.911 - 85.249) | 75.14 (74.915 - 75.371) |
|  | **PyTorch_EHR** | 85.22 (84.550 - 85.888) | **78.33% (77.267 - 79.384)*** |
|  | **PyTorch_EHR**  **Pre-Trained** | - | **81.14% (80.506-81.767)**** |
| Pneumonia | **LR** | 79.86% (79.851- 79.859) | 71.85% (71.849 - 71.852) |
|  | **LGBM** | 82.931% (82.735 - 83.094) | 75.59% (75.33 - 75.854) |
|  | **PyTorch_EHR** | **84.12% (83.435 - 84.813)*** | 75.93% (75.161 - 75.707) |
|  | **PyTorch_EHR**  **Pre-Trained** | - | 75.96% (75.192 - 76.736) |
| Skin Soft Tissue Infections | **LR** | 75.65% (75.648 - 75.658) | 75.96% (75.963 - 75.965) |
|  | **LGBM** | 78.88% (78.765 - 78.990) | 75.96% (75.963 - 75.965) |
|  | **PyTorch_EHR** | **80.06% (79.685 -80.444)*** | **80.62% (80.223 - 81.019)*** |
|  | **PyTorch_EHR**  **Pre-Trained** | - | **79.93 (79.552 - 80.317)** |

1. S Figure5: Features contributing to the individual prediction in an example patient

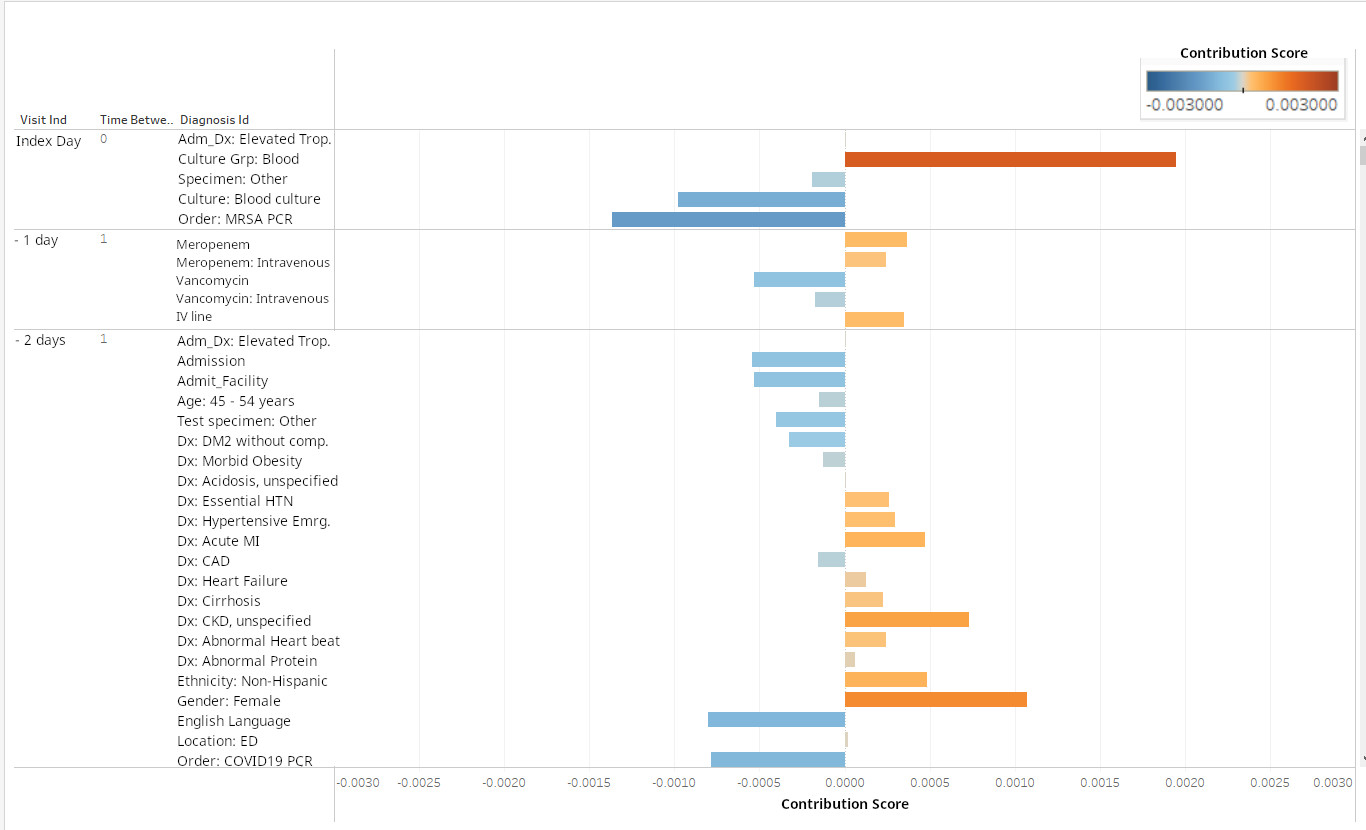

This patient was a female patient aged between 45 – 54 years with multiple underlying multiple comorbidities listed on admission two days (-2 days) before the index culture (blood culture on index date). Our model identified the risk score 0.541. After patient was admitted to the hospital, she started vancomycin and meropenem and blood culture was ordered. Subsequently cultures identified MRSA over two weeks.
